# Relationship between astrocyte reactivity, using novel ^11^C-BU99008 PET, and glucose metabolism, grey matter volume and amyloid load in cognitively impaired individuals

**DOI:** 10.1101/2021.08.10.21261690

**Authors:** Nicholas R Livingston, Valeria Calsolaro, Rainer Hinz, Joseph Nowell, Sanara Raza, Steve Gentleman, Robin J Tyacke, Jim Myers, Ashwin V Venkataraman, Robert Perneczky, Roger N Gunn, Eugenii A Rabiner, Christine A Parker, Philip S Murphy, Paul B Wren, David J Nutt, Paul M Matthews, Paul Edison

## Abstract

*Post mortem* neuropathology suggests that astrocyte reactivity may play a significant role in neurodegeneration in Alzheimer’s disease. We explored this *in vivo* using multimodal PET and MRI imaging. Twenty subjects (11 older, cognitively impaired subjects and 9 age-matched healthy controls) underwent brain scanning using the novel reactive astrocyte PET tracer ^11^C-BU99008, ^18^F-FDG and ^18^F-florbetaben PET, and T1-weighted MRI. Differences between cognitively impaired subjects and healthy controls in voxel-wise levels of astrocyte reactivity, glucose metabolism and grey matter volume were explored, and their relationship to each other was assessed using Biological Parametric Mapping (BPM). Aβ-positive cognitively impaired subjects showed greater brain astrocyte reactivity, except in the temporal lobe, with further increased astrocyte reactivity in Mild Cognitive Impairment compared to Alzheimer’s subjects in the cingulate cortices. BPM correlations revealed regions which showed reduced ^11^C-BU99008 uptake in Aβ-positive cognitively impaired subjects, such as the temporal lobe, also showed reduced ^18^F-FDG uptake and grey matter volume. BPM analysis also revealed a regionally-dynamic relationship between astrocyte reactivity and amyloid uptake: increased amyloid load in cortical association areas of the temporal lobe and cingulate cortices was associated with *reduced* astrocyte reactivity, whilst increased amyloid uptake in primary motor and sensory areas (in which amyloid load occurs later) was associated with *increased* astrocyte reactivity. These novel observations add to the hypothesis that while astrocyte reactivity may be triggered by early Aβ-deposition, sustained pro-inflammatory astrocyte reactivity with greater amyloid deposition may lead to astrocyte dystrophy and amyloid-associated neuropathology such as grey matter atrophy and glucose hypometabolism.

## 1. Introduction

Astrocytes are integral to normal brain function, playing important roles in neurogenesis, synaptogenesis, control of blood-brain barrier permeability and maintaining extracellular homeostasis^1^. In Alzheimer’s disease (AD), astrocytes can assume a reactive phenotype in which they become hypertrophic with the upregulation of glial fibrillary acidic protein (GFAP)^2^. Astrocyte reactivity associated with amyloid beta (Aβ) plaques is believed to have a neuroprotective role in pre-symptomatic and early AD^3^ through expression of proteases involved in the enzymatic cleavage and removal of Aβ^4^. However, with higher levels of Aβ, astrocyte reactivity can produce neurotoxic reactive oxygen species and inflammatory cytokines^5^. Astrocytes in AD also can lose normal neuroprotective capabilities as they become dystrophic with the progression of AD pathology^6, 7^.

Glucose hypometabolism, measured using ^18^F-fluorodeoxyglucose (^18^F-FDG) PET, and brain atrophy, measured using MRI, were two of the earliest neuroimaging markers of neurodegeneration developed for AD^8^. By contributing to synaptic loss and neurodegeneration, pro-inflammatory and dystrophic astroglia may be associated with accelerated grey matter atrophy^9^. Astrocytes are also necessary for metabolic support of neuronal activity^10^, so AD related changes in astrocytes might contribute directly to the brain glucose hypometabolism characteristic of AD^8^.

The novel PET tracer ^11^C-BU99008 has high specificity and selectivity for binding sites of type-2 imidazoline receptors (I_2_-BS), which are expressed primarily within astrocytes and are up-regulated with reactivity^11^. This tracer thus allows the study of astrocyte reactivity *in vivo*^12-17^. Pathologically increased ^11^C-BU99008 PET signal recently has been demonstrated in neurodegenerative disorders including AD^18^ and Parkinson’s disease^19^. Currently, the only available PET tracer which can measure astrocyte reactivity *in vivo* is ^11^C-deuterium-_L_-deprenyl (^11^C-DED)^20, 21^. However, this tracer binds to monoamine oxidase-B (MAO-B), which is not significantly elevated in late stage Aβ-deposition. The increased sensitivity of ^11^C-BU99008 over ^11^C-DED to detect astrocyte reactivity has recently been demonstrated in a preclinical model of AD^22^, thus warranting it’s use through further study in this clinical population. The aim of this study was to evaluate the relationship between astrocyte reactivity, using ^11^C-BU99008 PET, glucose metabolism, grey matter atrophy and Aβ-deposition in cognitively impaired subjects with a clinical diagnosis of AD-related dementia or mild cognitive impairment (MCI).

## 2. Materials and Methods

We recruited 20 subjects for this pilot study. Ethical approval for this study was obtained from the local and regional Research Ethics Committee, whilst approval to administer radiotracers was obtained from the Administration of Radioactive Substances Advisory Committee (ARSAC) UK. The human biological samples sourced from participants were obtained ethically and their research use was in accordance with the terms of the informed consent.

### 2.1 Subjects

Subjects were recruited from memory clinics, research registries and advertisements. We included 11 cognitively impaired subjects with a clinical diagnosis of AD-related dementia or MCI (6 AD, 5 MCI; Mini-Mental Status Examination (MMSE) score [mean ±SD] = 22.6 ±4.1) and 9 age-matched healthy volunteers without a history of brain disease (MMSE score [mean ±SD] = 29.1 ±1.27). The inclusion criteria for cognitively impaired subjects included the ability to give informed consent, an MMSE score ≥17 and at least 8 years of education. Exclusion criteria included contradictions to MRI and any evidence of significant small vessel or vascular disease on MRI. All subjects underwent medical and detailed cognitive assessments using the Repeatable Battery for the Assessment of Neuropsychological Status (RBANS), as well as ^11^C-BU99008, ^18^F-FDG and ^18^F-florbetaben PET and T1-weighted structural MRI. Aβ-positivity was defined by using a whole brain uptake cut-off of 1.43^23^.

### 2.2 Image acquisition

All image acquisition was performed at the Invicro Centre for Imaging Sciences in London, UK. MRI images were acquired using either a 3 Tesla Magnetom Trio or Verio (Siemens Healthcare Sector, Erlangen, Germany) with a 32-receiver channel head matrix coil. All PET imaging was performed on a Siemens Truepoint PET/CT scanner.

#### 2.2.1 MRI

#### 2.2.1 Structural MRI

All subjects underwent a sagittal T1-weighted MPRAGE, acquired with TR=2400 ms, TE=3.06 ms, flip angle=9°, TI=900 ms, matrix=[256 x 246], a 1 mm isotropic voxel size, anteroposterior phase encoding direction, IPAT factor 2 and a symmetric echo.

#### 2.2.2 PET

##### 2.2.2.1 ^11^C-BU99008 PET

All subjects underwent ^11^C-BU99008 PET scanning to assess astrocyte reactivity in the brain. ^11^C-BU99008 was synthesised on site. An initial CT scan was acquired for attenuation correction of the PET images, before a mean activity of 330 (±30) MBq ^11^C-BU99008 in 20ml normal saline was injected into the antecubital vein. Dynamic emission ^11^C-BU99008 PET images were acquired over 120 minutes and rebinned into 29 timeframes: 8×15s, 3×60s, 5×120s, 5×300s, and 8×600s. All subjects had arterial blood sampled continuously for the first 15 minutes, with twelve additional samples taken at 5, 10, 15, 20, 25, 30, 40, 50, 60, 70, 80, and 100 minutes after injection. A gamma counter was used to measure radioactivity in the whole blood and plasma for each sample. Reverse-phase high-performance liquid chromatography was used to evaluate metabolism of ^11^C-BU99008 by calculating the relative proportions of parent tracer and metabolites in the blood. Parametric images (Impulse Response Function at 120 minutes (IRF-120)) of ^11^C-BU99008 was generated using spectral analysis. This was performed using MICK-PM (Modelling, Input Functions and Compartmental Kinetics Parametric Map) software (available on request from Wolfson Molecular Imaging Centre, University of Manchester, Manchester, UK).

##### 2.2.2.2 ^18^F-FDG and ^18^F-florbetaben PET

All subjects also underwent ^18^F-FDG and ^18^F-florbetaben PET scanning to assess glucose metabolism and Aβ-deposition in the brain, respectively. Subjects received a target dose of 185 MBq ^18^F-FDG and 236.4 (±6.8) MBq ^18^F-florbetaben as single intravenous boluses in the respective scanning sessions. For ^18^F-FDG scans, PET acquisition commenced 30 minutes after tracer injection, and the scans were acquired for 30 minutes. Using MICKPM, activity over the last 30 minutes was averaged, resulting in a 3D 30-60min ^18^F-FDG add image. For ^18^F-florbetaben scans, PET acquisition commenced 90 minutes after tracer administration and the subjects were scanned for 30 minutes. Activity over the 30-minute acquisition period was averaged, resulting in a 3D 90-120min ^18^F-florbetaben add image.

### 2.3 Image processing

MRI and PET images were pre-processed using SPM12 (Wellcome centre for human neuroimaging, UCL, London, UK) in MATLAB (v2014a). 3D PET data was co-registered to the structural MRI of each subject. The structural MRI was segmented into grey matter (GM), white matter (WM) and CSF, and the GM and WM maps were used to generate a study-specific template using diffeomorphic anatomical registration through exponentiated lie algebra (DARTEL)^24^. The DARTEL flow fields were then used to normalise each of the coregistered PET images and GM maps to MNI space and an 8mm FWHM Gaussian kernel was used to smooth the data. Tracer uptake for ^11^C-BU99008 PET was calculated through spectral analysis (IRF-120min). Tracer uptake for ^18^F-FDG and ^18^F-florbetaben PET was evaluated using the standardised uptake value ratio (SUVR) and the Hammers atlas^25^, referenced to the pons grey & white matter and the cerebellar grey matter, respectively. This was done by dividing the cerebral cortical ^18^F-FDG and ^18^F-florbetaben mean images by the uptake value of the relevant reference region, which had been calculated in Analyze 11.0 (developed by the Biomedical Imaging Resource (BIR) at the mayo clinic). This resulted in smoothed normalised ^18^F-FDG, ^18^F-florbetaben, ^11^C-BU99008 and GM voxel-based morphometry (VBM) images that were used to assess glucose metabolism, Aβ deposition, astrocyte reactivity and grey matter atrophy patterns, respectively. This was done through voxel-wise statistical parametric mapping (SPM) and biological parametric mapping (BPM) analysis.

### 2.4 SPM analysis

Voxel-level SPM analysis was performed in order to better characterise the spatial distribution of tracer uptake difference between the cognitively impaired subjects and the healthy controls. The smoothed normalised ^11^C-BU99008 IRF-120 parametric maps, 30-60min ^18^F-FDG add images, 90-120min ^18^F-florbetaben add images and VBM GM images of all subjects were entered into 4 separate two-sample Student’s t-test in SPM12 (2-tailed).

### 2.5 BPM and ROI correlation analysis

In order to assess the neuroanatomical relationship between ^11^C-BU99008 binding and glucose metabolism, Aβ deposition and GM atrophy, BPM and ROI correlation analysis was performed. For the ROI correlations, subject-specific object maps were created from the Hammers atlas^25, 26^ and were used to sample the ROI radioactivity concentration for the 3 normalised (not smoothed) PET images, as well as the ROI volume of the VBM images. The ROIs included the frontal lobe, temporal lobe, medial temporal lobe, parietal lobe, occipital lobe, anterior cingulate, posterior cingulate and the whole brain (made up of the 4 lobes and the cingulate). Correlation between each of the 4 imaging measures in each of the four lobes and whole brain for Aβ-positive patients was calculated using Pearson’s correlation coefficient in SPSS (v26, released 2019). For the BPM correlations, Z-score maps for each of the 4 imaging modalities were created. These represent tracer uptake and GM atrophy patterns relative to the healthy control’s mean and standard deviation for each subject on a voxel-level basis, calculated with the following formulae

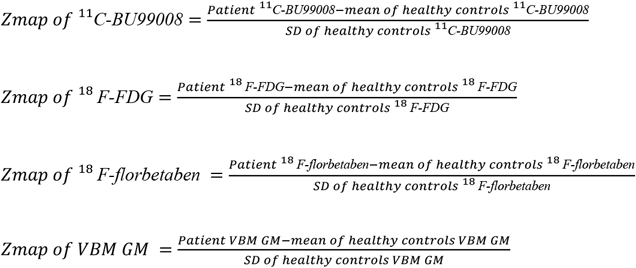

Voxel-level correlations between ^11^C-BU99008 and the remaining three modalities were estimated for all patients using BPM^27^, an SPM toolbox that runs through MATLAB. Additional correlations were run on Aβ-positive patients between ^11^C-BU99008 and ^18^F-FDG & VBM GM.

## 3. Results

All the healthy controls were Aβ-negative, 7 of the patients were Aβ-positive (4 AD, 3 MCI) and 4 patients were Aβ-negative (2 AD, 2 MCI).

### 3.1 Group-level SPM analysis

Two-sample t-tests in SPM contrasting Aβ-positive patients and healthy controls showed distributions of differences in tracer uptake and grey matter volumes that were consistent with the ROI-analyses. Aβ-positive patients had increased ^11^C-BU99008 uptake particularly in the frontal and occipital lobes (Figure 1a), reduced ^18^F-FDG uptake in the temporal, parietal and occipital lobes (Figure 1b), reduced grey matter volume in temporal regions, particularly the hippocampi (Figure 1c), and increased ^18^F-florbetaben uptake in frontotemporal regions (Figure 1d). An exploratory two-sample t-test comparing MCI and AD subjects showed increased ^11^C-BU99008 uptake in MCI patients, particularly in the frontal and temporal regions.

**Figure 1:**
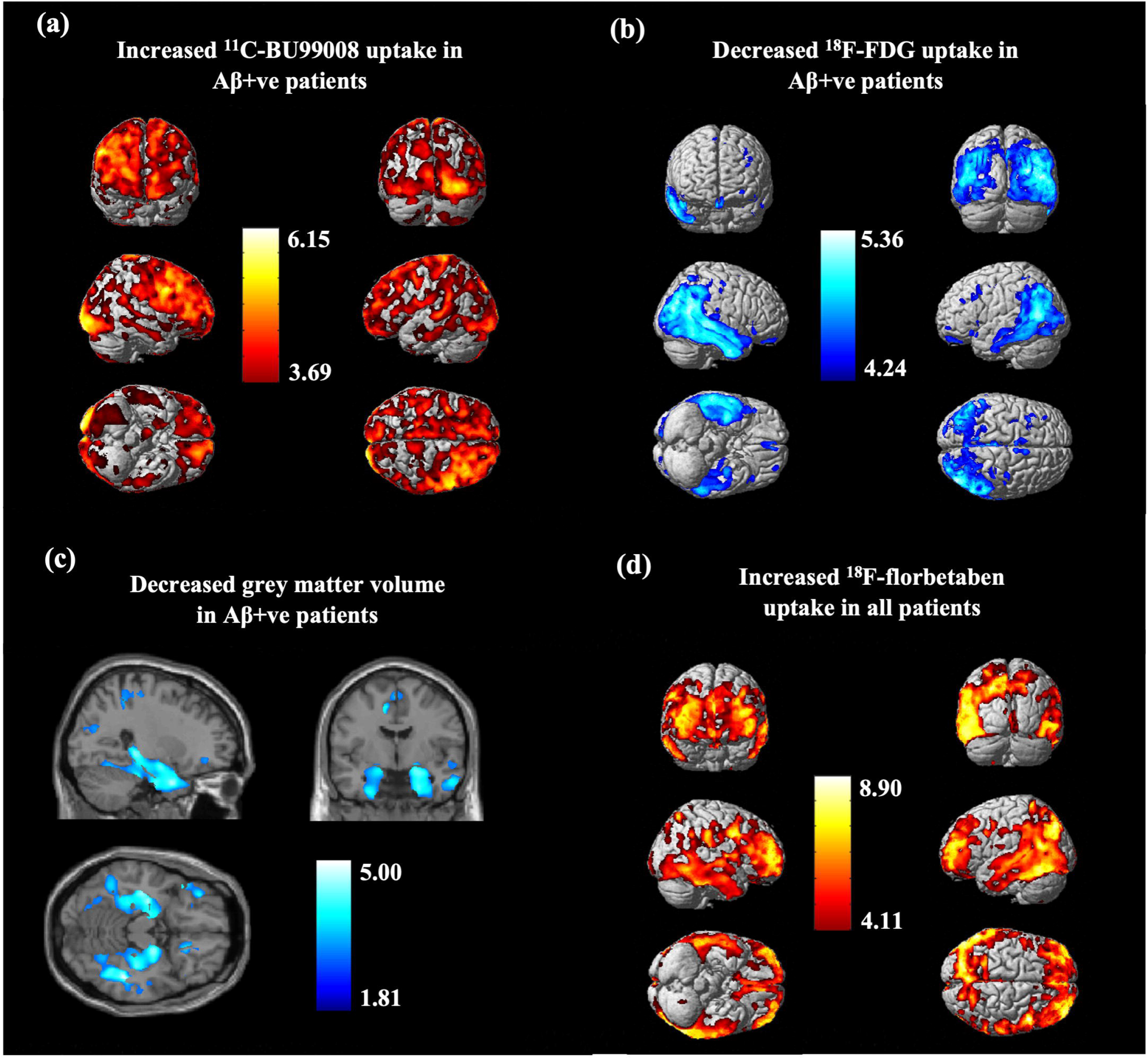
Statistical Parametric Mapping (SPM) group analysis in patients compared to healthy controls. **(a**) Increased ^11^C-BU99008 uptake in Aβ-positive patients compared to healthy controls, rendered at cluster threshold of p<0.05 and an extent threshold of 50 voxels. **(b)** Decreased ^18^F-FDG uptake in Aβ-positive patients compared to health controls, rendered at cluster threshold of p<0.001 and an extent threshold of 50 voxels. **(c)** Decreased grey matter volume in Aβ-positive patients compared to healthy controls, rendered at cluster threshold of p<0.05 and an extent threshold of 50 voxels. **(d)** Increased ^18^F-Florbetaben uptake in all patients compared to healthy controls, rendered at cluster threshold of p<0.001 and an extent threshold of 50 voxels. Colourbar units are contrast estimates.

### 3.2 Regional and Voxel-wise Correlations

#### 3.2.1 ^11^C-BU99008 x ^18^F-FDG

BPM analysis showed that reduced ^11^C-BU99008 uptake was correlated with reduced ^18^F-FDG uptake, particularly in the temporal, parietal and frontal lobes. ROI correlations suggested the same directions for correlations, although none of the regions reached statistical significance frontal (r=0.202, p=0.664), occipital (r=0.264, p=0.567), temporal (r=0.567, p=0.184) and parietal (r=0.622, p=0.135) lobes, and the whole brain (r=0.478, p=0.279).

#### 3.2.2 ^11^C-BU99008 x VBM

BPM analysis showed reduced ^11^C-BU99008 uptake was correlated with reduced grey matter volume in the frontal and temporal lobes. ROI correlations showed the same correlation, showing strong correlations in the frontal (r=0.808, p=0.028), temporal (r=0.935, p=0.002), parietal (r=0.833, p=0.020) and occipital lobes (r=0.762, p=0.047), as well as the whole brain (r=0.901, p=0.006).

#### 3.2.3 ^11^C-BU99008 x ^18^F-florbetaben

BPM analysis described an inverse correlation of increased ^18^F-florbetaben uptake with reduced ^11^C-BU99008 uptake in regions such as the temporal lobe and the cingulate, whilst increased ^18^F-florbetaben uptake was positively correlated with increased ^11^C-BU99008 uptake in primary motor and primary sensory areas. ROI analyses showed that reduced ^11^C-BU99008 uptake was correlated with increased ^18^F-florbetaben uptake, particularly in the frontal (r=-0.780, p=0.039), temporal (r=-0.779, p=0.039), occipital (r=-0.911, p=0.004) lobe and the whole brain (r=-0.798, p=0.032).

## 4. Discussion

In this study, we used the novel imidazoline receptor PET tracer ^11^C-BU99008 to test for evidence of a dynamic relationship between astrocyte reactivity and amyloid-associated neurodegeneration based on tissue hypometabolism and atrophy measured using ^18^F-FDG PET and structural MRI, respectively. We found evidence for increased astrocyte reactivity in Aβ-positive patients, primarily in frontal, parietal and occipital regions. These increases were greater in MCI than AD patients. Regional correlational analyses showed that lower astrocyte reactivity in Aβ-positive patients was associated with both glucose hypometabolism in the parietal, temporal and frontal lobes and grey matter atrophy in frontal and temporal lobes. However, analyses of regional differences in the relationships between PET markers of Aβ-deposition and astrocyte reactivity displayed a striking heterogeneity; greater Aβ-deposition was associated with increased astrocyte reactivity in primary motor and primary sensory cortical areas, but decreased astrocyte reactivity in temporal regions.

^11^C-BU99008 is a novel PET tracer that binds to I_2_-BS, expression of which is associated with astrocyte reactivity^28, 29^. Brain I_2_-BS is upregulated with healthy aging^30^, and is further increased in AD^31^. The sensitivity and specificity of ^11^C-BU99008 to bind to I_2_-BS expressing reactive astrocytes has been further evidenced in a recent autoradiography study of AD brains where tracer uptake was greater compared to cognitively normal brains^22^. In line with this, we found cognitively impaired subjects showed increased ^11^C-BU99008 uptake compared to healthy controls, corroborating earlier studies with another PET marker of astrocyte reactivity, ^11^C-deuterium-L-deprenyl (^11^C-DED)^32^. Additionally, we found increased ^11^C-BU99008 uptake in MCI subjects compared to AD subjects, particularly in the frontal lobe. Interestingly, another ^11^C-DED study also found increased uptake in the frontal lobe in Aβ-positive MCI, but not AD, subjects compared to healthy controls^20^. Both these findings agree with the hypothesis that astrocyte reactivity is an early event in the progression of AD pathology, occurring in response to early amyloid deposition, which typically originates in the frontal lobe^33^. In the early stages, reactive astrocytes have a neuroprotective role, aiding in the clearance of Aβ^4^. Further evidence of this hypothesis comes from a subsequent study that showed increased ^11^C-DED binding in autosomal dominant AD patients early in their disease progression^34^, primarily in temporal regions, another region involved in early Aβ deposition^35^. In our cohort of Aβ-positive MCI and AD subjects, we found lower ^11^C-BU99008 binding in the temporal lobe, which was associated with greater relative progression of amyloid-associated neuropathology, that is glucose hypometabolism and grey matter atrophy. We propose this reduced ^11^C-BU99008 uptake in the temporal lobe region reflects astrocyte dystrophy^36^, brought about by an amyloid-induced chronic pro-inflammatory and neurotoxic astrocyte phenotype^37^ and resulting in reduced glycolytic capacity and secondary impaired neuronal metabolism^38^ or cell loss^39, 40^. Correlations between regional reductions in ^11^C-DED and^18^F-FDG PET signals similar to those described here also were associated with regionally more advanced ^11^C-PIB PET pathology in a longitudinal study of people with autosomal dominant AD or MCI^41^. Interaction of Aβ with reactive astrocytes has been proposed as a trigger for astrocytes to switch from a neuroprotective to a neurotoxic role.

There are obvious limitations to our study. First, only a small number of subjects were able to be imaged. While this is a pilot study, the explanatory power was enhanced by the design in which uptake of the three PET tracers and brain volume all were assessed in the same people. A second limitation was the cross-sectional design, which we acknowledge; however, *post mortem* pathology has the same limitation. Our results thus are better interpreted descriptively and as suggestive of a hypothetical model, rather than a strong, independent test. Nonetheless, the consistency of directions of effect observed in this study and the earlier ^11^C-DED PET studies^20, 41^ provides compelling support for the model proposed. That is, astrocyte reactivity occurs in response to early Aβ-deposition, aiding in the clearance of Aβ, but following interactions with high levels of Aβ the astrocytes become neurotoxic, contributing to reduced tissue activity and cell death that is associated with cognitive impairment. It also strengthens confidence in the earlier work, which otherwise suffers from uncertainties regarding the specificity of binding of ^11^C-DED in the brain^20^. Nonetheless, ^11^C-BU99008 can detect astrocyte reactivity with a greater sensitivity than ^11^C-DED^22^, especially amongst higher levels of amyloid load^18, 42^, and thus should be prioritised.

In conclusion, this study supports neuropathological observations arguing that astrocyte reactivity with amyloid-related neuropathology is dynamic^2^. We have demonstrated *in vivo* with the novel PET tracer ^11^C-BU99008 that astrocyte reactivity is increased in regions presumed to represent earlier stages of pathological progression with low Aβ-deposition loads, and conversely relatively reduced in regions that show signs of more advanced disease progression with greater Aβ-deposition and atrophy. In the absence of molecular imaging markers intrinsically discriminating different microglial activation phenotypes, our multi-modal imaging approach may allow relevant inferences to be made from the relative ^11^C-BU99008, ^18^F-FDG and ^18^F-florbetaben PET signals and brain volume sensitive MRI measures. Future, larger, longitudinal studies are needed to further test this dynamic model and, if supported, interventions developed to arrest progression of the neurotoxic phenotypic transformation of astrocytes in AD.

## Data Availability

All the necessary data is presented in the manuscript.

## Acknowledgements

The authors thank Invicro Centre for Imaging Sciences for the provision of ^11^C-BU99008, scanning and blood analysis equipment. The authors also thank Piramal Life Sciences/Life Molecular Imaging for providing the ^18^F-florbetaben and permission to acquire unlabelled florbetaben. We thank Dementia Platform UK (DPUK) and GSK for the generous funding for this project. This research was co-funded by the NIHR Imperial Biomedical Research Centre and was supported by the NIHR Imperial Clinical Research Facility. The views expressed are those of the authors and not necessarily those of NHS, the NIHR nor the Department of Health. P.E. was funded by the Medical Research Council and now by Higher Education Funding Council for England (HEFCE). He has also received grants from Alzheimer’s Research, UK, Alzheimer’s Drug Discovery Foundation, Alzheimer’s Society, UK, Alzheimer’s association, US, Medical Research Council, UK, Novo Nordisk, Piramal Life Sciences and GE Healthcare. P.M.M. gratefully acknowledges generous support from Edmond J Safra Foundation and Lily Safra, the NIHR Investigator programme and the UK Dementia Research Institute.

